# Statistical Analysis Plan for the manuscript reporting the primary outcome of the MSU-TELEMED study

**DOI:** 10.1101/2025.02.11.25322103

**Authors:** Leonid Churilov, Hannah Johns, Vignan Yogendrakumar, Anna Balabanski, MSU-TELEMED Trial Investigators

## Abstract

This manuscript describes the Statistical Analysis Plan (SAP) for the following study: Evaluating the Safety and Efficacy of Telemedicine Physician Assessments on a Mobile Stroke Unit: a Prospective Open-Label Blinded End-Point Randomized Controlled Trial (MSU-TELEMED)

This document is a supplement to the MSU-TELEMED study protocol and contains the statistical analysis plan (SAP) for the main paper of the trial in which the primary outcome will be reported. This document complies with the guidelines for the content of statistical analysis plans in clinical trials

## 1 Abbreviations

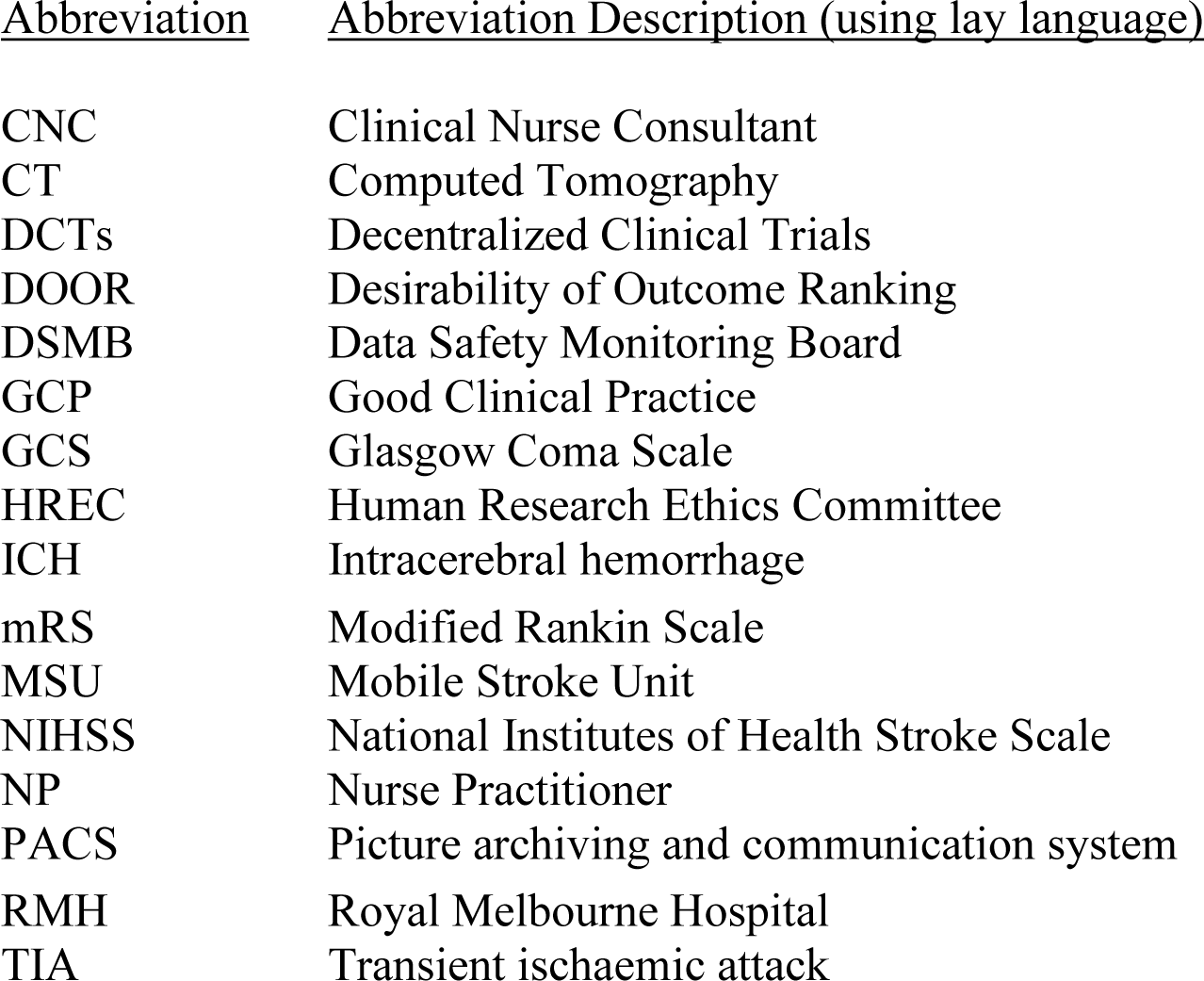

| <u>Abbreviation</u> | <u>Abbreviation Description (using lay language)</u> |
| --- | --- |
| CNC | Clinical Nurse Consultant |
| CT | Computed Tomography |
| DCTs | Decentralized Clinical Trials |
| DOOR | Desirability of Outcome Ranking |
| DSMB | Data Safety Monitoring Board |
| GCP | Good Clinical Practice |
| GCS | Glasgow Coma Scale |
| HREC | Human Research Ethics Committee |
| ICH | Intracerebral hemorrhage |
| mRS | Modified Rankin Scale |
| MSU | Mobile Stroke Unit |
| NIHSS | National Institutes of Health Stroke Scale |
| NP | Nurse Practitioner |
| PACS | Picture archiving and communication system |
| RMH | Royal Melbourne Hospital |
| TIA | Transient ischaemic attack |

### 2 Administrative Information

Protocol: 2023.110; Version 3.0; 19 Sep 2024 Clinicaltrials.gov register Identifier: NCT05991310

### 2.1 Document Version History

| Version Date | Version | Author | Signature | Change Description | Reason/Comment |
| --- | --- | --- | --- | --- | --- |
| 12 Dec 2024 | 1.0 | Leonid Churilov | Redacted in publicly available version | Initial release | Not applicable |
|  |  | Hannah Johns | Redacted in publicly available version |  |  |

### 2.2 Approvals

The undersigned have reviewed this plan and approve it as final. They find it to be consistent with the requirements of the protocol as it applies to their respective areas. They also find it to be compliant with ICH-E9 principles and confirm that this analysis plan was developed in a completely blinded manner (i.e., without knowledge of the effect of the intervention being assessed)

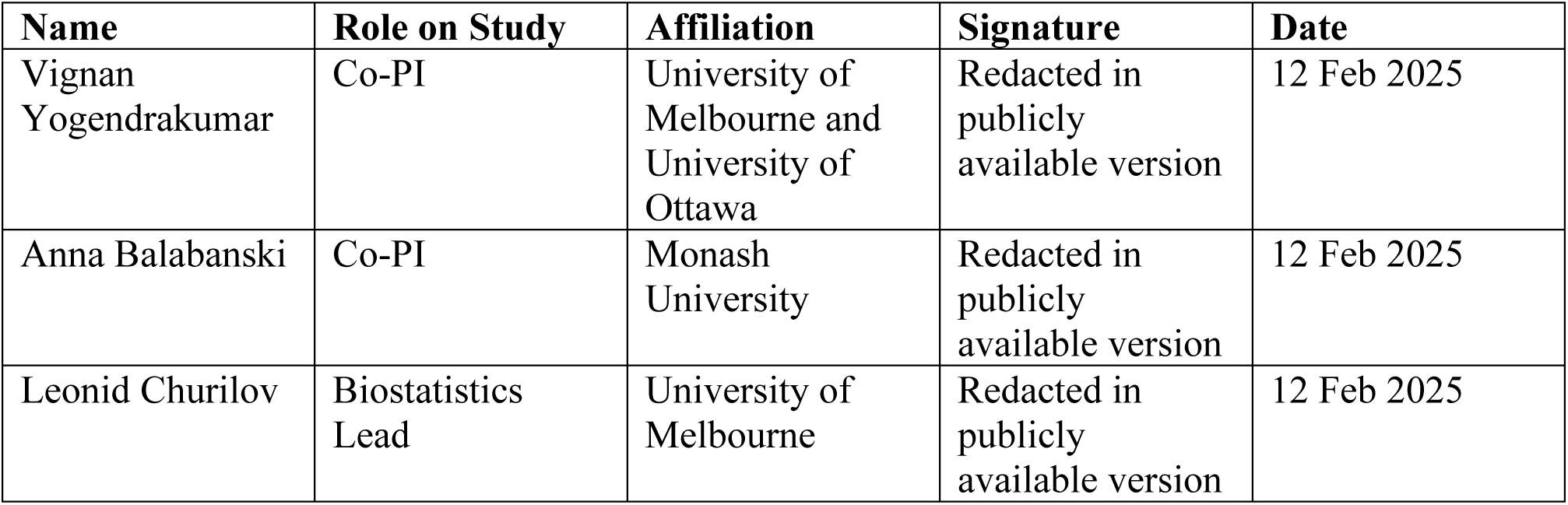

| Name | Role on Study | Affiliation | Signature | Date |
| --- | --- | --- | --- | --- |
| Vignan Yogendrakumar | Co-PI | University of Melbourne and University of Ottawa | Redacted in publicly available version | 12 Feb 2025 |
| Anna Balabanski | Co-PI | Monash University | Redacted in publicly available version | 12 Feb 2025 |
| Leonid Churilov | Biostatistics Lead | University of Melbourne | Redacted in publicly available version | 12 Feb 2025 |

## 3 Study design

### 3.1 Overview

Mobile Stroke Units (MSU; ’stroke ambulance’) are ambulances with built-in brain imaging devices staffed by a team of paramedics and stroke specialists, including a stroke neurologist, nurse and radiographer, who are able to provide time-critical stroke care in the prehospital setting. MSUs have been shown to provide patient care resulting in improved clinical outcomes and faster access to time-critical stroke treatments, without compromising patient safety.

However, the costs associated with establishing and staffing of MSUs in Australia currently limit their use to densely populated cities.

The MSU-TELEMED trial is a prospective, randomized, open-label blinded end-point parallel-arm, single-center MSU trial comparing 2 staffing models: (1) a neurologist review of patients via telemedicine (intervention) versus (2) an onboard neurologist assessing patients in person (comparator). The Melbourne MSU has operated within Melbourne, Victoria, since 2017 and is Australia’s first MSU. Based at the Royal Melbourne Hospital, a large comprehensive stroke center, the MSU operates within a 20-km radius and services a population of 1.7 million people within metropolitan Melbourne, a city of over 5 million people. Due to funding and staffing constraints, it currently operates 10 hours per day on weekdays.

### **2.2** Aims and hypotheses

The primary aim of MSU-TELEMED Trial is to test the hypothesis that the telemedicine neurologist model will provide superior resource efficiency without compromising safety or timely delivery of care, when compared with an onboard model of care.

### **2.3** Patient population

#### Inclusion criteria

- adult patients aged ≥18 years
- presenting within 24 hours of symptom onset or last known well
- who undergo assessment by the Melbourne MSU without cancellation before the MSU attending the patient

#### Exclusion criteria

- attendance of the Melbourne MSU is deemed unnecessary by either the local paramedic team or the MSU team on the basis of provided information before arrival on-scene,
- the patient presents significant medical or logistical challenges that greatly delay standard treatment, or
- any other medical contraindication at the discretion of the investigator

### **2.4** Randomization

#### Overview

The days the neurologist is onboard or on telemedicine are randomized in a covariate-adaptive manner. Randomization is undertaken by units of days and is balanced for the level of nursing expertise, defined as Nurse Practitioner versus Clinical Nurse Consultant level of training. An adaptive component is used to balance the number of patients reviewed by each nurse type with an onboard neurologist in person and telemedicine arms. Nurse Practitioners are nurses with extensive experience in stroke who have undergone formal training (e.g., a master’s degree) with a specific focus in acute, subacute, and chronic stroke care. In contrast, Clinical Nurse Consultants are nurses who may come from varying backgrounds (emergency, neurology, general practice) and are trained to manage stroke care specific to the MSU.

#### Technical Details

The onboard or telemedicine days are randomized one week in advance to minimize staffing disruption for the MSU. We sought to balance the number of participants that would be seen by nurses with different training (Nurse Practitioner vs Comprehensive Nurse Consultant) during randomization. However, due to the need to randomize ahead of time, anticipated last-minute changes to service (e.g. nursing staff change due to illness, the MSU going out of service due to breakdown) and an unknown number of MSU patients that would be seen on each day, conventional randomization algorithms that balance participant-level covariates (e.g. biased-coin minimization or stratified permuted blocks) could not be immediately applied.

After exploring potential modifications to existing randomization algorithms, we implemented a modified version of Pocock biased coin minimization [1] to allocate neurologists to be onboard or telemedicine on a daily level, accounting for the expected operation and staff schedule at time of randomization and the anticipated number of participants on each day. Pocock minimization was chosen because it was the most straightforward to modify and would automatically compensate for last-minute schedule changes at the next randomization schedule.

The modified adaptive randomization algorithm was as follows:

1. Estimate the expected number of participants on each day based on accrued data (calculated as the total number of participants seen divided by the number of days the trial has been active). If no participants have been enrolled yet, assume 1 participant per day.
2. Cross-tabulate the number of participants seen by each type of nurse
3. For each day in this week, starting with Monday:

a. Randomly allocate a neurologist to be onboard or via telemedicine using Pocock biased coin (strength 0.7) randomization to balance the expected type of nurse using the cross-tabulation;
b. Add the expected number of participants to the cell of the cross-tabulation matching the expected nurse type and arm allocation from step 3a;
c. Continue to the next day to be randomized, using the updated cross-tabulation.

The modified adaptive randomization algorithm assumes that there was no seasonality or other temporal trends that needed to be considered when estimating the expected number of participants on each day. Following clinical input from, and discussions with, the MSU-TELEMED Management Committee, this assumption was considered reasonable.

The adaptive randomization schedules were generated in a rolling fashion weekly by Co-PI Yogendrakumar using a custom-built R shiny app.

### 3.5 Baseline and follow-up assessments

All responsible investigators receive training in the systems for data collection and data entry and training in Good Clinical Practice (GCP). All investigators responsible for patient assessment participate in training in critical assessment items (NIHSS, mRS).

The assessment schedule for this trial is found in the protocol. The baseline assessments include NIHSS, pre-morbid mRS, demographic data, past medical history, previous stroke history and cerebral imaging information.

Safety endpoints are reviewed and adjudicated centrally by two medical monitors who are blinded to treatment allocation. Patients who are actively treated on the MSU (administration of thrombolysis and/or transfer for endovascular therapy) are followed at 24hr and at 3 months unless death occurs. The assessment at 24 hours is an evaluation of post-treatment imaging for hemorrhagic transformation, performed centrally by an assessor who is blinded to treatment allocation. The 3-month assessments of post-stroke disability (mRS) are conducted by hospital staff where the patient was admitted, via assessors who are blind to treatment allocation.

The trial management team are responsible for ensuring that all data are completed in a timely manner. Patients do not receive payment for participation.

### 3.6 Sample size considerations

As discussed in detail in the Definitions of the Outcomes and Estimands section of this SAP, the primary outcome for this study is the odds that a randomly selected participant in the Telemedicine arm will have a better outcome than a randomly selected participant in the onboard neurologist arm are measured using the hierarchical outcomes approach, including, in order of importance: (1) safety, (2) scene-to-decision-time metrics, and (3) resource efficiency.

We hypothesized that the Telemedicine intervention will result in a distribution of win/tie/loss proportions of 0.5/0.2/0.3 that corresponds to the win odds (0.5 + 0.2/2)/(0.3 + 0.2/2) = 0.6/0.4 = 1.5 when ties are split equally between treatment arms.

This effect estimate is conservative due to efficiency likely being substantially greater in the Telemedicine arm and was deemed adequate to compensate for a small potentially arising design effect inflation due to the cluster-randomized nature of the day-, rather than individual participant-based, randomization. Appropriate a priori design effect estimation was not deemed feasible due to the lack of prior data on within- vs between-day Intraclass Correlation Coefficient (ICC) on MSU. At the same time, due to the nature of acute stroke, relatively low expected cluster size (low counts of eligible MSU patients per day), and no existing evidence on the effect of day of the week on the type of stroke patients presenting on a given day, both the within-day clustering effect and the resulting design effect due to cluster randomization was deemed to be small.

Recruiting 242 participants (121 per arm) would yield a power of 0.8 to observe such or higher hypothesized treatment effect against the null hypothesis of no treatment effect (win odds = 1) assuming a 2-sided α level of 0.05. [2] To account for 10% of potential losses due to non-evaluable data, the final sample size for the study is set at 270 participants.

The last participant was recruited on 08 August 2024, resulting in the total achieved sample size of 275 participants.

### 3.7 Unblinding

Only the Data and Safety Monitoring Board (DSMB) have access to interim data and results. The DSMB reviewed unblinded data in accordance with the DSMB Charter. Treatment allocations are securely stored and separated from the assessors involved in safety assessments, symptomatic intracranial hemorrhage, and 3-month mRS. Statisticians not involved in the DSMB remain blinded and work on dummy datasets until the computer codes for statistical analysis are validated.

### 3.8 Definitions of Primary Estimand and Secondary and Exploratory Outcomes

This trial follows the Estimand framework for the design and analysis of clinical trials. The objective of the Estimand framework as presented in addendum (R1) to the International Council on Harmonization E9 guidance [3], is to align the clinical trial objectives with the study design, endpoints and analysis, in order to improve study planning and the interpretation of the study results.

The Estimand framework requires appropriate pre-specification of population, individual-level outcome measure, population level summary measure, and strategies for handling intercurrent events [4]. The trial Primary Estimand and secondary and exploratory outcomes are described below while respective analysis methods are detailed in Statistical Analysis section of this SAP.

#### Primary Estimand: safe and timely delivery of care with superior resource efficiency

**Population:** All included participants as reflected by the trial inclusion and exclusion criteria

**Individual-level outcome measure:** the hierarchical composite endpoint of safe and timely delivery of care with superior resource efficiency as defined in SAP Table 1.

**Population-level summary outcome measure:** the Win Odds as described in the Statistical Analysis Section. The Win Odds greater than 1 is indicative of the Telemedicine arm being more likely to experience the better outcomes of interest; a Win Odds less than 1 is indicative of a less favorable effect in the Telemedicine arm as compared to the doctor onboard arm.

**Strategy for Intercurrent events:** *Treatment policy strategy* - any intercurrent events which prevent collection of the primary outcome measure according to the rules specified in SAP Table 1 will be treated according to a treatment policy strategy. Missingness mechanism will be examined for randomness and standard strategies for dealing with missing data will be employed as specified in the Statistical Analysis section.

This definition of the Estimand adheres to the intention to treat (ITT) principle and will be complemented by the ITT trial analysis.

**SAP Table 1:** Individual outcome measure for the Primary Estimand.

| **SAP Table 1: Individual outcome measure for the Primary Estimand** |  |
| --- | --- |
| Hierarchical composite outcome involving, in order of importance: |  |
| 1. Safety<br>2. Scene-to-decision-time metrics<br>3. Resource usage |  |
| where individual components are defined as follows: |  |
| Safety metrics | 1. Clinician unable to complete the assessment<br>2. Post-thrombolysis complications (occurrence, identification, and management): Angioedema; symptomatic intracerebral hemorrhage after thrombolysis; major extracranial bleeding after thrombolysis<br>3. Deterioration during assessment: National Institutes of Health Stroke Scale increase by $\geq 4$, Glasgow Coma Scale decrease by $\geq 2$, airway compromise, breathing compromise, circulatory compromise, vomiting, seizure |
|  | A participant will be classified as having a negative safety outcome if they experience at least one of the above end points.<br>Having a negative safety outcome is classified as a loss compared with not having such an outcome for the purpose of the primary outcome analysis |
| Scene-to-decision-time metrics | On-scene to definitive decision making, including:<br>a. Decision for thrombolysis<br>b. Decision to refer for EVT<br>c. Decision to stand down case<br>d. Decision to transfer to stroke hospital (eg, suspected stroke but unable to treat on MSU) |
| Resource usage | The percentage of “productive time” the neurologist was actively involved in the care of the patient.<br>This is defined as:<br>a) Onboard model: the time the neurologist is providing clinical care divided by the time from MSU dispatch to the time the MSU has returned to base or to the next dispatch<br>b) Telemedicine model: the time the neurologist is providing clinical care divided by the time from the first call from the MSU to the neurologist to the time to the end of the last interaction with the MSU |

#### Secondary Safety Estimand

**Population:** All included participants as reflected by the trial inclusion and exclusion criteria

**Individual-level outcome measure:** presence or absence of safety events as defined in SAP Table 1

**Population-level summary outcome measure:** proportion of participants with safety events

**Strategy for Intercurrent events:** *Treatment policy strategy* - any intercurrent events which prevent collection of the outcome measure will be treated according to a treatment policy strategy. Missingness mechanism will be examined for randomness and standard strategies for dealing with missing data will be employed as specified in the Statistical Analysis section.

#### Secondary Time-to-Decision Estimand

**Population:** All included participants for whom definitive treatment decision has been made

**Individual-level outcome measure:** Time from arrival on-scene to definitive treatment decision, min;

**Population-level summary outcome measure:** Median time from arrival on-scene to definitive treatment decision, min;

**Strategy for Intercurrent events:** *Principal stratum strategy* – time to decision will be assessed in participants who did not experience intercurrent events preventing a definitive treatment decision such as clinician’s inability to complete assessment due to, e.g., a technical or telecommunication failure.

#### Secondary Neurologist “Percentage of Productive Time” Estimand

**Population:** All included participants as reflected by the trial inclusion and exclusion criteria

**Individual-level outcome measure:** Percentage of neurologist “productive” time as defined in SAP Table 1.

**Population-level summary outcome measure:** Median percentage of neurologist “productive” time

**Strategy for Intercurrent events:** *Treatment policy strategy* - any intercurrent events which prevent collection of the outcome measure will be treated according to a treatment policy strategy. Missingness mechanism will be examined for randomness and standard strategies for dealing with missing data will be employed as specified in the Statistical Analysis section.

#### Exploratory Safety, Process, and Functional Outcomes

Safety: Deterioration during assessment:

• Proportion of participants with NIHSS increase by 4 points or more

• Proportion of participants with GCS decrease by 2 points or more

• Proportion of participants with airway and breathing compromise

• Proportion of participants with circulatory compromise

• Proportion of participants vomiting

• Proportion of participants experiencing a seizure

Safety: Post-thrombolytic complications:

• Proportion of participants with angioedema

• Proportion of participants with Symptomatic ICH

• Proportion of participants with major extracranial bleeding

Safety: Technical Issues

• Proportion of cases where clinician is unable to complete assessment

• Proportion of cases with telemedicine failures

• Proportion of cases with imaging acquisition failures or delays

• Proportion of cases with other technical issues

Treatment Decisions:

• Proportion of cases where CT and/or CTA was performed

• Proportion of cases with thrombolytic administered and/or transfer for endovascular thrombectomy

• Proportion of cases where transfer to stroke centre is arranged and no on-scene treatment is provided

• Proportion of cases with stand-down and transfer of care to local ambulance crew

Time metrics:

• Proportion of cases with time from imaging performed to imaging (non-contrast CT) reviewed below 5 mins, 5-10 mins, and above 10 mins

• Time from imaging performed to imaging (CT angiogram) reviewed below 5 mins, 5-10 mins, and above 10 mins

• Time from arrival on-scene to thrombolytic administration, min

• Time from MSU dispatch to arrival on-scene, min

• Time from arrival on-scene to neurologist activation, min

• Time from arrival on-scene to imaging assessment (non-contrast CT), min

Functional outcomes:

• Modified Rankin Scale at 90 days post stroke

## 5 Statistical analysis

### 5.1 Analysis principles and general considerations

- All outcomes and analyses are prospectively categorized as primary, secondary, or exploratory
- Differences in all endpoints between the two arms of the study will be tested independently at the two-tailed 0.05 level of significance. Unless specifically stated otherwise, all estimates of treatment effects will be presented with 95% confidence intervals (95%CIs)
- No formal adjustments will be undertaken to constrain the overall type I error associated with the secondary, tertiary, and exploratory analyses. Their purpose is to supplement evidence from the primary analysis to more fully characterize the treatment effect. Results from the secondary and exploratory analyses will be interpreted in this context.
- Analysis for primary outcome and all secondary outcomes analyses will be conducted will be conducted within the Estimand framework using the Estimands described earlier in this SAP while preserving the principle that all participants will be analyzed within the arm to which they were randomized, utilizing pre-specified strategies for handling intercurrent events.

The analysis strategy for MSU-TELEMED trial is defined as follows [5]:

- It is based on a design that aims to collect all outcome data on all randomized subjects
- It includes a main analysis that keeps subjects in their randomized arms, analyzes all available outcome data, and is valid under a named plausible assumption about the missing data (data missing at random)
- It includes sensitivity analyses that consider a range of plausible alternative assumptions that contradict the main assumption about the missing data
- All individuals are included in sensitivity analyses

Subgroup analyses could be carried out irrespective of whether there is a significant treatment effect on the outcome. Their purpose is to supplement evidence from the primary analysis to help to fully characterize the treatment effect. Results from subgroup analyses will be interpreted in this context. The following subgroups are prespecified as being of interest for the primary analysis:

- Age: <75 year vs >=75 years
- Sex: Male vs Female
- Nurse Type: Nurse Practitioners vs Clinical Nurse Consultants
- Baseline NIHSS: 0-4 vs >=5
- Use of imaging: CT and/or CTA vs no CT/CTA imaging
- MSU Diagnosis: Ischemic or hemorrhagic stroke or TIA vs. other
- Analyses will be conducted using Stata and R statistical software.

### 5.2 Interim analyses

The Independent DSMB reviewed unblinded data. No formal interim analyses for efficacy or futility were planned or conducted.

### 5.3 Trial profile

Flow of patients through the study will be displayed in a standard CONSORT diagram. The report will include the number of patients included, withdrawn, lost to follow up, the number who received the allocated treatment and were analyzed.

### 5.4 Patients’ characteristics and baseline comparisons

In order to assess balance, description of the specified baseline characteristics will be presented for the Telemedicine and Onboard arms (Section 7 of this SAP). Discrete variables will be summarized as frequencies and percentages. Unless otherwise indicated in the tables, percentages will be calculated according to the number of patients for whom data are available. If there are more than 5% missing values, the denominator will be added in a footnote in the corresponding summary table. Continuous variables will be summarized by use of either mean and standard deviation (SD) or median and interquartile range (IQR). Durations and time intervals will be summarized by medians and IQRs.

### 5.5 Primary outcome: safe and timely delivery of care with superior resource efficiency

#### Outcome measure

The odds of the Telemedicine arm having better outcomes than the onboard neurologist arm using the hierarchical outcomes approach, including, in order of importance: (1) safety, (2) scene-to-decision-time metrics, and (3) resource efficiency.

To estimate such odds (referred to as Win Odds), each participant in the Telemedicine arm will be compared to every patient in the onboard assessment arm to produce a “win”, “loss” or “tie” using the following sequential decision-making process will be performed (SAP Figure 1): Safety assessment: If a participant in one treatment arm is achieving better safety than the comparator as listed in SAP Table 1, this is defined as a “win” for that participant and a “loss” for the comparator. Having a negative safety outcome is classified as a loss compared with not having such an outcome for the purpose of the primary outcome comparison. If there is no difference in safety, the second step is undertaken, where the time-to-treatment decision is compared.

1. Time-to-treatment metrics assessment: If the time-to-treatment decision for a participant in one treatment arm is greater than the respective time for a participant in the comparator arm by more than a predetermined clinically meaningful difference of 15 minutes, this is defined as a “loss” for that participant and a “win” for the comparator. We selected a clinically meaningful time difference of 15 minutes on the basis of prior pilot evaluations of MSU telemedicine models elsewhere. If there is no clinically meaningful difference in the time-to-treatment decision, then the third step, resource usage (the “productive percentage” of neurologist’s time), is compared.
2. Resource usage assessment: If a participant’s management is associated with a >10% greater percentage of productive time than the comparator as described in SAP Table 1, this is defined as a win for that participant and a loss for the comparator. If there is no difference in resource usage, the 2 participants are declared as tied for the overall outcome.

**SAP Figure 1** (adopted from [6]):

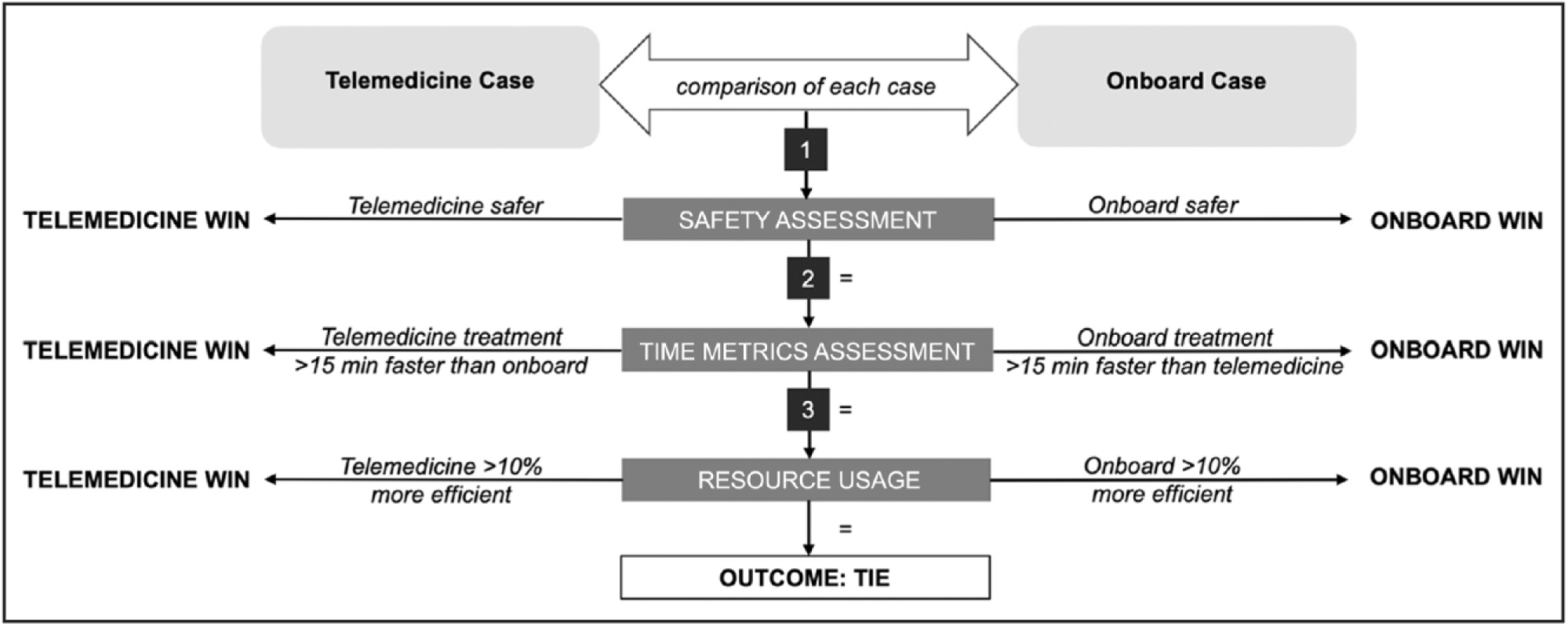

Subsequently, to obtain the estimate of Win Odds the total number of wins plus half of the total number of ties are divided by the total number of losses plus half of the total number of ties and to obtain the estimate of Net Benefit the total number of losses is subtracted from the total number of wins.

#### Statistical Hypothesis

The null hypothesis to be rejected is that the Win Odds is 1.

#### Treatment of missing values

Based on blinded monitoring of the amount of missing primary outcome data in the MSU- TELEMED trial, it is anticipated that there will be minimal missing primary outcome data. Nevertheless, a small amount of missing data may be inevitable.

Following clinical input from, and discussions with, the MSU-TELEMED Management Committee, the individual components of the hierarchical primary outcome are assumed to be missing-at-random (MAR), i.e. it is assumed that the probability of the outcome data being missing may depend on the values of the observed data, but not on the values of the missing data. Important explanatory and auxiliary variables were collected and will be examined to assess the plausibility of the MAR assumption.

For the primary analysis, missing primary outcome data and covariates required for stratification will be imputed using multiple imputations with chain equations (10 imputations, 30 chains, 5000 bootstrap iterations) with the subsequent use of Rubin’s rules.

Sensitivity analyses using the Tipping Point method [7,8] will be conducted.

#### Analysis method

This analysis will be stratified by baseline NIHSS (0-4 vs >=5) and whether a CT scan was performed or not in a full-factorial manner. Pooled estimates will be obtained through Mantel-Haenszel style weights [9] applied to these strata.

Hypothesis testing for the primary analysis will be performed via rerandomization tests as per FDA guidelines [10]. Participants will be repeatedly rerandomized into Telemedicine and Neurologist onboard arms with all outcomes remaining fixed to ensure that the null hypothesis is true. The Win Odds and Net Benefit will be estimated under each rerandomization and compared to the observed treatment effect. The p-value will be estimated as the proportion of times that a rerandomized win odds is higher than the observed treatment effect.

The treatment effect will be presented as stratified Win Odds and Net Benefit with the corresponding 95% CIs. Confidence intervals for primary analysis will be estimated through bootstrap resampling. To account for clustering within days, we will randomly resample days with replacement, selecting all participants within each day without replacement, until the number of days match those in the trial. This approach, rather than fixing the number of participants, best mimics variation properties driven by cluster effects [11].

10,000 replications will be used for both the rerandomization test and resampling. If the Win Odds and Net Benefit are not estimable for any of these replications (due to e.g. all participants in one stratum being allocated to a single arm upon resampling or rerandomization), further replications will be performed in blocks of 5,000 until at least 10,000 successful replications are completed.

Excess replications will be discarded from estimation of treatment effects. The proportion of total replications attempted that were not successful will be reported.

### 5.6 Secondary safety outcome

#### Outcome measure

Proportion of participants with safety events (as defined by Secondary Safety Estimand)

#### Statistical Hypothesis

The set of statistical hypotheses is p(Telemedicine)=p(Neurologist Onboard) versus p(Telemedicine) ≠ p(Neurologist Onboard), where p(Telemedicine) is the proportion of participants with at least one safety event in the Telemedicine arm and p(Neurologist Onboard) is the proportion of participants with at least one safety event in the Neurologist Onboard arm.

#### Treatment of missing values

As for the Primary Outcome.

#### Analysis method

The treatment effect will be presented as stratified risk differences and stratified risk ratios with the corresponding 95% CIs. The analysis will follow the procedure described above for the Primary Outcome.

### 5.7 Secondary Time-to-Decision Outcome

#### Outcome measure

Median time from arrival on-scene to definitive treatment decision in minutes

#### Statistical Hypothesis

The null hypothesis to be refuted is that in participants for whom the definitive treatment decision has been made, the median time from arrival on-scene to definitive treatment decision in the Telemedicine arm is equal to that in the Neurologist Onboard arm.

#### Treatment of missing values

As for the Primary Outcome restricted to the participants for whom the definitive treatment decision has been made.

#### Analysis method

The treatment effect will be presented as difference in medians with corresponding 95% Cis, adjusted for stratification variables used in the Primary Outcome. We will also report respective effect sizes and 95%CIs for the between-arm differences in the 25^th^ and the 75^th^ percentile of the time distributions. The analysis will follow the procedure described above for the Primary Outcome, using quantile regression models for each rerandomization.

### 5.8 Secondary Neurologist “Percentage of Productive Time” outcome

#### Outcome measure

Median percentage of neurologist “productive” time (as defined by the Secondary Neurologist “Percentage of Productive Time” Estimand).

#### Statistical Hypothesis

The null hypothesis to be refuted is that the median neurologist productive time in the Telemedicine arm is equal to that in the Neurologist Onboard arm.

#### Treatment of missing values

As for the Primary Outcome.

#### Analysis method

The treatment effect will be presented as difference in medians with corresponding 95% CIs, adjusted for stratification variables used in the Primary Outcome. We will also report respective effect sizes and 95%CIs for the between-arm differences in the 25^th^ and the 75^th^ percentile of the percentage of productive time distributions. The analysis will follow the procedure described above for the Primary Outcome, using quantile regression models for each rerandomization.

### 5.9 Exploratory outcomes (some or all of these outcomes may be reported in publications subsequent to primary paper)

Analyses for all the exploratory outcomes will be conducted with the aim to estimate the magnitude of respective effect sizes and related uncertainty in the form of the 95%CI. Effect sizes will be presented as stratified/adjusted risk differences and risk ratios for the pre-specified dichotomous and categorical outcomes; between-arm differences in the median, 25^th^ percentile and 75^th^ percentile of respective distributions for the pre-specified time metrics; and generalized or common odds ratio for the ordinal analysis of the modified Rankin Scale.

Exploratory analyses of the diagnostic utility of stroke MSU diagnosis using the final diagnosis at discharge as the reference standard will be conducted using Receiver Operating Characteristic (ROC) curves. Area under the ROC, sensitivity, specificity, as well as Cohen’s kappa for statistical agreement will be separately reported for two individual arms of the study.

### 5.10 Tables and figures for the main paper

The proposed tables and figures for the main results are presented in Appendix 1. Manuscript Table 1 will report key baseline characteristics of participants by treatment arm.

Manuscript Table 2 will report the primary and secondary outcomes. Manuscript Supplementary tables S1 and S2 will report various exploratory safety and process outcomes as well as diagnostic utility of stroke diagnosis on MSU by treatment arm. Manuscript Supplementary Table S3 will provide the full listing of adverse events.

Figure 1 will be the CONSORT diagram. Figure 2 will present the Win Odds Plot for the primary hierarchical composite outcome. Figure 3 will present the scatter plot of every paired treatment-to-control participant comparison with the x-axis representing the difference in time to treatment decision between the two participants and the y-axis representing the difference in the percentage of the neurologist productive time. Supplementary Figure S1 will present Forest plots for the pre-specified subgroup analyses for the primary and secondary outcomes Supplementary Figure S2 will report “Grotta bar” stacked bar chart of each grade on the mRS in each treatment arm for the participants who received thrombolysis or endovascular treatment by treatment arm.

## Data Availability

All data produced in the present work are contained in the manuscript

## 5 Manuscript Tables and Figures

Figure 1: Consort Diagram

Figure 2: Primary Outcome (Win Odds Plot)

Figure 3: Difference in time to definitive treatment decision vs difference in percentage of neurologist productive time scatter plot

Figure S1: Forest plots for the pre-specified subgroup analyses for the primary and secondary outcomes

Figure S2: mRS (“Grotta”) bars for those who received EVT or thrombolysis by treatment arm

**Manuscript Table 1:** Baseline Participant Characteristics (n= )

|  | Telemedicine<br>(n=) | Neurologist<br>Onboard<br>(n=) |
| --- | --- | --- |
| Age, years; median (IQR) |  |  |
| Female Sex |  |  |
| Stroke Nurse Status |  |  |
| Nurse Practitioner |  |  |
| Clinical Nurse Consultant |  |  |
| Medical History |  |  |
| Hypertension |  |  |
| Diabetes |  |  |
| Elevated Cholesterol |  |  |
| Atrial Fibrillation |  |  |
| Prior Ischemic Stroke or Transient Ischemic Attack |  |  |
| Pre-stroke score on the mRS |  |  |
| 0 |  |  |
| 1 |  |  |
| 2 |  |  |
| 3 |  |  |
| 4 |  |  |
| 5 |  |  |
| Clinical Information |  |  |
| GCS; median (IQR) |  |  |
| NIHSS; median (IQR) |  |  |
| Antiplatelet Use |  |  |
| Anticoagulant Use |  |  |
| Provisional MSU Diagnosis |  |  |
| Ischemic Stroke |  |  |
| Transient Ischemic Attack |  |  |
| Intracerebral Hemorrhage |  |  |
| Subarachnoid/Subdural Hemorrhage |  |  |
| Seizure |  |  |
| Migraine |  |  |
| Other |  |  |
| Symptom onset |  |  |
| Witnessed |  |  |
| Wake-up |  |  |
| LKW unknown |  |  |

Results are (n,%) unless otherwise stated; Abbreviations: IQR=interquartile range; NIHSS=National Institutes of Health Stroke Scale; GCS=Glasgow coma scale; mRS=modified Rankin scale; LKW=last known well

**Manuscript Table 2:** Primary and Secondary Outcomes.

|  | **Telemedicine**<br>**(n=)** | **Neurologist**<br>**Onboard (n=)** | **Effect Size (95%**<br>**CI)** |
| --- | --- | --- | --- |
| Primary Outcome | Number/Proportion of Telemedicine Wins<br>Number/Proportion of Ties<br>Number/Proportion of Neurologist Onboard Wins |  | Stratified Win Odds<br>Stratified Net Benefit |
| Secondary Outcomes |  |  |  |
| Safety Events (n,%) |  |  | Stratified Risk Difference<br>Stratified Risk Ratio |
| Time from arrival on-scene to definitive treatment decision, min; median (IQR) |  |  | Adjusted Difference in:<br>25<sup>th</sup> percentile<br>Median<br>75<sup>th</sup> percentile |
| Percentage of neurologist “productive” time; median (IQR) |  |  | Adjusted Difference in:<br>25<sup>th</sup> percentile<br>Median<br>75<sup>th</sup> percentile |

Results are (n,%) unless otherwise stated
Abbreviations: IQR=interquartile range; NIHSS=National Institutes of Health Stroke Scale; GCS=Glasgow coma scale; mRS=modified Rankin scale; LKW=last known well

**Manuscript Supplementary Table 1:** Safety and Process Outcomes.

|  | Telemedicine<br>(n=) | Neurologist<br>Onboard (n=) | Effect Size<br>(95% CI) |
| --- | --- | --- | --- |
| **Safety** |  |  |  |
| Deterioration during assessment |  |  |  |
| NIHSS increase by 4 or greater |  |  |  |
| GCS decrease by 2 or greater |  |  |  |
| Airway and breathing compromise |  |  |  |
| Circulatory Compromise |  |  |  |
| Vomiting |  |  |  |
| Seizure |  |  |  |
| Post-thrombolytic complications |  |  |  |
| Symptomatic ICH |  |  |  |
| Major extracranial bleeding |  |  |  |
| Angioedema |  |  |  |
| **Technical Issues** |  |  |  |
| Clinician unable to complete assessment |  |  |  |
| Telemedicine failures |  |  |  |
| Imaging acquisition failures or delays |  |  |  |
| Other |  |  |  |
| **Treatment Decisions** |  |  |  |
| Perform CT and/or CTA |  |  |  |
| Administer thrombolytic and/or transfer for endovascular thrombectomy |  |  |  |
| Arrange transfer to stroke center (no on-scene treatment provided, transfer performed by local ambulance team) |  |  |  |
| Stand-down and transition to local ambulance team |  |  |  |
| **Time Metrics** |  |  |  |
| Time from imaging performed to imaging (non-contrast CT) reviewed |  |  |  |
| < 5 min |  |  |  |
| 5-10 min |  |  |  |
| > 10 min |  |  |  |
| Time from imaging performed to imaging (CT angiogram) reviewed |  |  |  |
| < 5 min |  |  |  |
| 5-10 min |  |  |  |
| > 10 min |  |  |  |
| Time from MSU dispatch to arrival on-Scene, min; median (IQR) |  |  |  |
| Time from arrival on-scene to neurologist activation, min; median (IQR) |  |  |  |
| Time from arrival on-scene to imaging assessment (non-contrast CT), min; median (IQR) |  |  |  |
| Time from arrival on-scene to imaging assessment (CT angiogram), min; median (IQR) |  |  |  |
| Time from arrival on-scene to thrombolytic administration, min; median (IQR) |  |  |  |

Results are (n,%) unless otherwise stated
Abbreviations: ICH=intracerebral hemorrhage; NIHSS=National Institutes of Health Stroke Scale; GCS=Glasgow coma scale;

**Manuscript Supplementary Table 2:** Stroke diagnosis on MSU vs at discharge.

| Telemedicine |  |  |  | Neurologist Onboard |  |  |  |
| --- | --- | --- | --- | --- | --- | --- | --- |
| MSU<br>Diagnosis | Final Diagnosis at Discharge |  |  | MSU<br>Diagnosis | Final Diagnosis at Discharge |  |  |
|  |  | Stroke | Non-Stroke |  |  | Stroke | Non-Stroke |
|  | Stroke |  |  |  | Stroke |  |  |
|  | Non-Stroke |  |  |  | Non-Stroke |  |  |
| Kappa |  |  |  | Kappa |  |  |  |
| AUC |  |  |  | AUC |  |  |  |
| Sensitivity |  |  |  | Sensitivity |  |  |  |
| Specificity |  |  |  | Specificity |  |  |  |

**Manuscript Supplementary Table 3:**
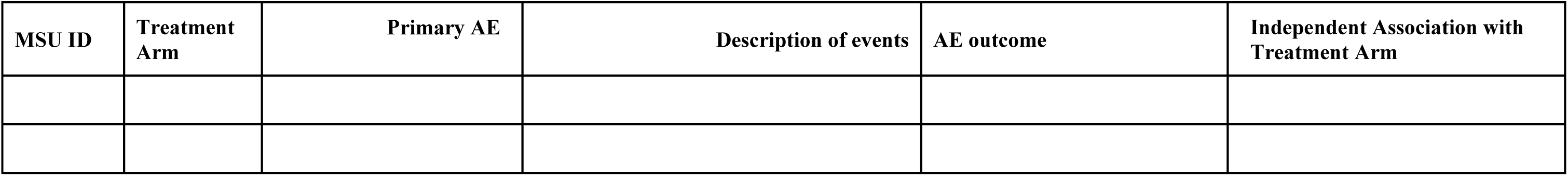
Full List of Adverse Events.

